# Outcome of COVID-19 with co-existing surgical emergencies in children: our initial experiences and recommendations

**DOI:** 10.1101/2020.08.01.20166371

**Authors:** Md Samiul Hasan, Md Ayub Ali, Umama Huq

**Affiliations:** Dhaka Shishu (Children) Hospital, Dhaka, Bangladesh; Ward – 6, Urology room, 2^nd^ Floor, Dhaka Shishu (Children) Hospital, Sher E Bangla Nagar, Dhaka- 1207, Bangladesh

**Keywords:** COVID 19, COVID 19 in children, Children’s Surgery, Surgical emergency, Surgery in COVID 19 positive patients

## Abstract

**Background:** COVID-19 has changed the practice of surgery vividly all over the world. Pediatric surgery is not an exception. Prioritization protocols allowing us to provide emergency surgical care to the children in need while controlling the pandemic spread. The aim of this study is to share our experiences with the outcome of children with COVID-19 who had a co-existing surgical emergency.

**Methods:** This is a retrospective observational study. We reviewed the epidemiological, clinical, and laboratory data of all patients admitted in our surgery department through the emergency department and later diagnosed to have COVID-19 by RT-PCR. The study duration was 3 months (April 2020 – June 2020). A nasopharyngeal swab was taken from all patients irrespective of symptoms to detect SARS CoV 2 by RT-PCR with the purpose of detecting asymptomatic patients and patients with atypical symptoms. Emergency surgical services were provided immediately without delay and patients with positive test results were isolated according to the hospital protocol. We divided the test positive patients into 4 age groups for the convenience of data analysis. Data were retrieved from hospital records and analyzed using SPSS (version 25) software. Ethical permission was taken from the hospital ethical review board.

**Results:** Total patients were 32. Seven (21.9%) of them were neonates. Twenty-four (75%) patients were male. The predominant diagnosis was acute abdomen followed by infantile hypertrophic pyloric stenosis (IHPS), myelomeningocele, and intussusception. Only two patients had mild respiratory symptoms (dry cough). Fever was present in 13 (40.6%) patients. Fourteen (43.8%) patients required surgical treatment. The mean duration of hospital stay was 5.5 days. One neonate with ARM died in the post-operative ward due to cardiac arrest. No patient had hypoxemia or organ failure. Seven health care workers (5.51%) including doctors & nurses got infected with SARS Co V2 during this period.

**Conclusion:** Our study has revealed a milder course of COVID-19 in children with minimal infectivity even when present in association with emergency surgical conditions. This might encourage a gradual restart to mitigate the impact of COVID-19 on children’s surgery.

## Introduction

COVID-19 storm is showing its lethal face around the world and it is still beyond prediction how long it will continue, and how deadly it could be. Increasingly more people are being affected and people with co-existing diseases are suffering more.[1] The causative agent severe acute respiratory syndrome coronavirus 2 (SARS CoV 2) spreading rapidly around the world. As of 23^rd^ July more than 15 million people got infected with SARS CoV 2 and more than 6 million people have lost their lives because of COVID-19.[2]

Fortunately, the infection rate in children is low in comparison to adult and most cases are mild or asymptomatic. Therefore, a detailed clinical picture of COVID-19 in children is yet to come. Moreover, presentations of COVID-19 in children are more inconsistent than adults. Only around 25% of children present with common respiratory symptoms which leads to delay in diagnosis & treatment and increases the risk of transmission.[3-6]

As an effort to control the rapid transmission, governments & organizations around the world have implemented distancing strategies. To comply with this strategy, healthcare institutions have taken several measures like discouraging hospital visits for non-emergency problems, prioritization, and re-scheduling health problems according to their time sensitivity. Pediatric surgical service is also not an exception. Different hospitals and associations have formalized protocols to ensure essential surgical service for children in need and at the same time to limit the spread of the virus and also to protect the health care workers.[7-10]

At the beginning of the outbreak of SARS CoV 2 in March in our country, we have also canceled all routine surgeries for children and been performing only emergency surgeries to limit the spread of the SARS CoV 2. The study aims to illustrate the outcomes we observed in children with COVID-19 who had co-existing surgical emergencies.

## Methods

This is a retrospective observational study. We reviewed the epidemiological, clinical, and laboratory data of all patients admitted in our surgery department through the emergency department and later diagnosed to have COVID-19 by RT-PCR. The study duration was 3 months (April 2020 – June 2020).

In this study period, only children with emergency symptoms were admitted. A nasopharyngeal swab was taken from all patients irrespective of symptoms to detect SARS CoV 2 by RT-PCR with the purpose of detecting asymptomatic patients and patients with atypical symptoms. Emergency surgical services were provided immediately without delay and patients with positive test results were isolated according to the hospital protocol. We divided the test positive patients into 4 age groups for the convenience of data analysis.

Data were retrieved from hospital records and analyzed using SPSS (version 25) software. Ethical permission was taken from the hospital ethical review board.

## Results

Total 32 children were tested positive for SARS CoV 2 by RT-PCR. Seven (21.9%) of them were neonates (Table 1). The smallest one tested positive was 3 days old. Twenty-four (75%) patients were male. The predominant diagnosis was acute abdomen followed by infantile hypertrophic pyloric stenosis (IHPS), myelomeningocele, and intussusception (Figure 1). Only 2 patients had mild respiratory symptoms (dry cough). Fever was present in 13 (40.6%) patients.[Supplemental material]

**Table 1:**
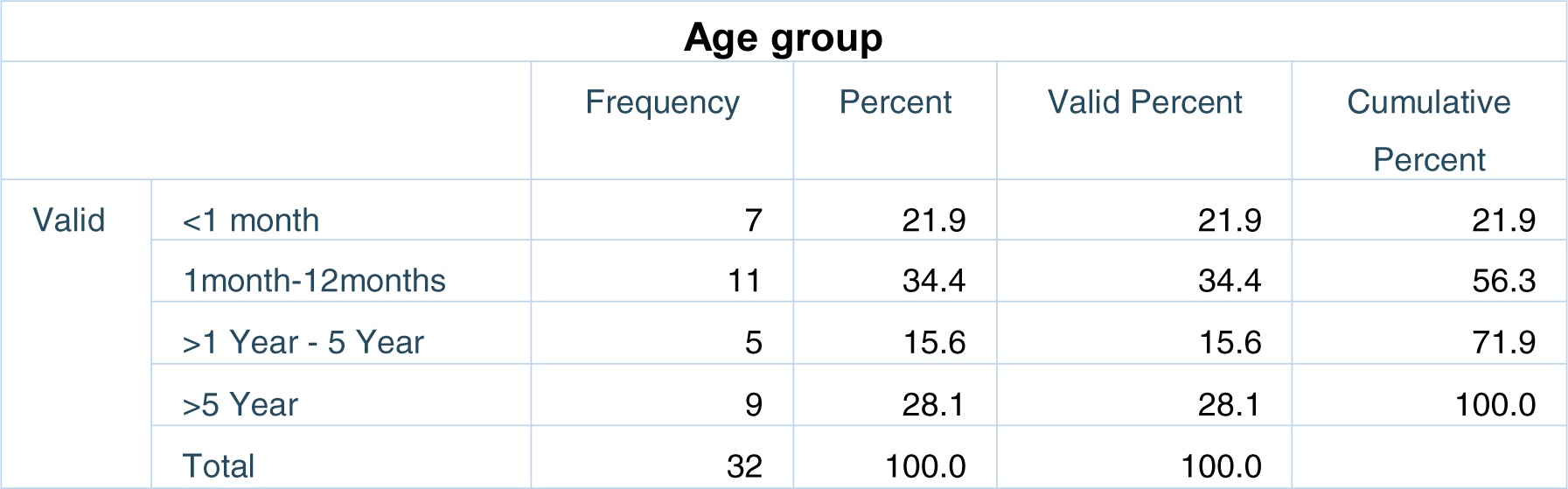
Age distribution of the patients.

**Figure 1:**
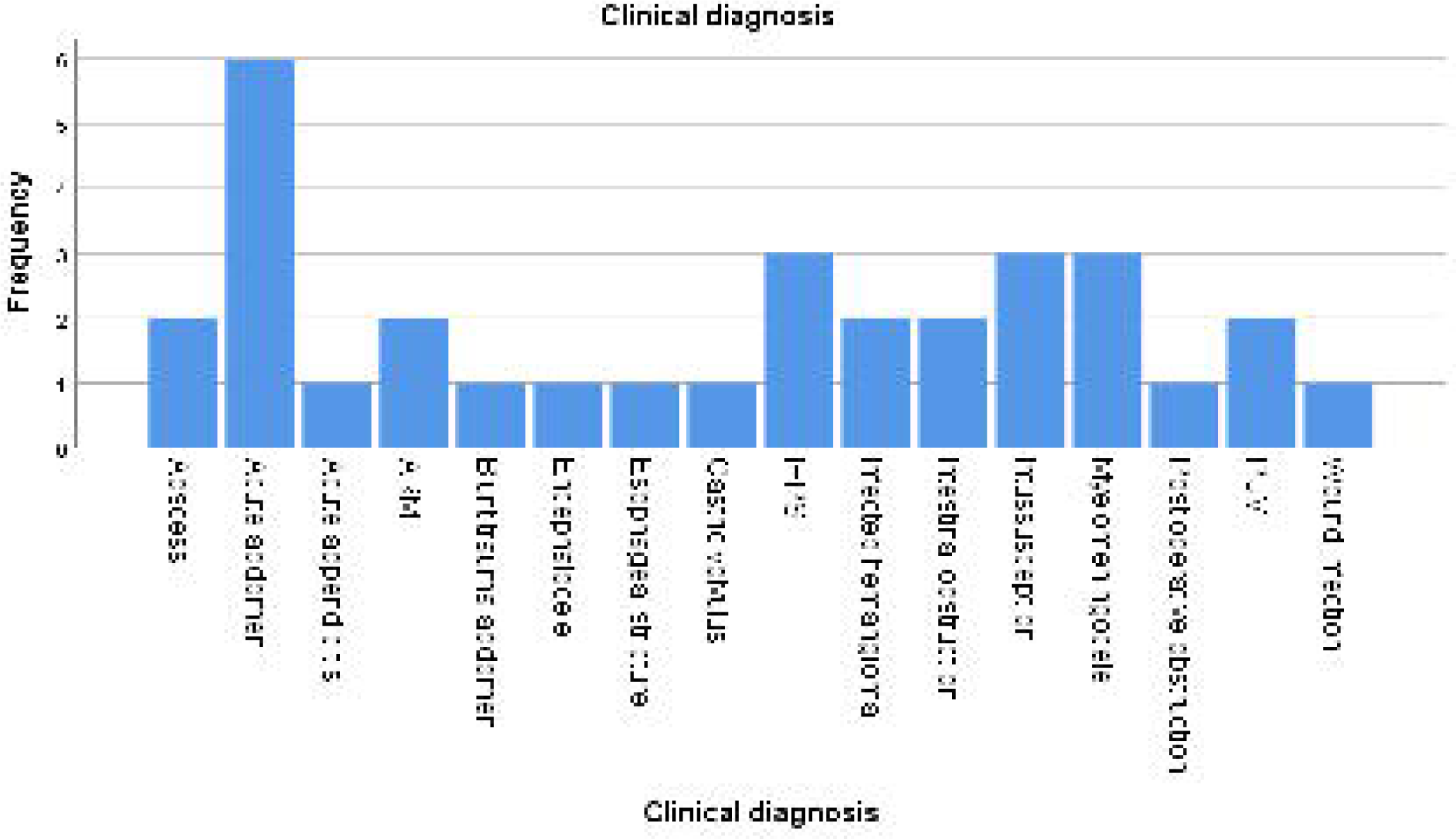
Clinical diagnosis of COVID-19 positive patients.

Apart from only one patient, none had any radiological evidence of COVID-19. Surgical treatment was provided to 14 (43.8%) patients (Table 2). The mean duration of hospital stay was 5.5±1.6 days.[Supplemental material] One neonate with anorectal malformation (ARM) died in post-operative ward due to cardiac arrest; other patients were discharged after managing surgical symptoms with advice for home isolation. No patient had hypoxemia or organ failure. Seven health care workers out of 127 (5.51%) including doctors & nurses got infected with SARS Co V2 during this period. All of them were mildly symptomatic, did not require hospital admission, and recovered smoothly.

**Table 2:** Modalities of treatment.

| Modality of treatment: Surgical or non-surgical |  |  |  |  |  |
| --- | --- | --- | --- | --- | --- |
|  |  | Frequency | Percent | Valid Percent | Cumulative Percent |
| Valid | Surgical | 14 | 43.8 | 43.8 | 43.8 |
|  | Non-surgical | 18 | 56.3 | 56.3 | 100.0 |
|  | Total | 32 | 100.0 | 100.0 |  |

## Discussion

COVID-19 has changed the way of surgical practice globally. This is a critical and unprecedented period, but efforts are being made to ensure inevitable emergency surgical care. The system is also adapting itself as more evidence is being generated.[7-11] From the beginning of the outbreak in our country, we have adopted the protocol to continue only the emergency admission at our department and try conservative treatment approaches.

More than 20 % of our patients were neonates. The mode of transmission in these patients was not transparent. This might be due to the parents’ tendency to hide history or reluctance to test when asymptomatic or mildly symptomatic. Vertical transmission of SARS CoV2 from mother to baby is not been proven yet. An increased level of virus-specific IgM has been reported in neonates of the infected mother, but this remains inconclusive due to the high rate of false positivity. Transmission through breast milk has also not been confirmed. Breast milk samples of mothers with COVID-19 pneumonia have been tested negative for SARS CoV 2.[12] These reports raise the suspicion of nosocomial transmission to neonates.

Two third of the children were male. It is not clear that the lower incidence of COVID-19 in women is also applicable to female children. Studies suggested that immune regulatory proteins encoded by the X chromosome cause less viral load and inflammation in females than male.[13]

The most common presentation was with gastrointestinal (GI) symptoms. Six patients presented with sudden onset of severe abdominal pain & several episodes of non-bilious vomiting. Laboratory investigations did not reveal any abnormality rather RT-PCR for SARS Co V2 came positive. One of them had radiological evidence of COVID-19 pneumonia in chest X-ray. No one had respiratory symptoms. All of them improved with conservative treatment. GI symptoms have been reported in up to 10% of COVID-19 patients.[14] A viral infection is the cause of around 50% of intussusceptions in children. COVID-19 has also been reported to cause intussusception in children, and pediatric surgeons in an epidemic area should be aware of this.[15-16]

Surgical treatment was required in 14 (43.8%) patients. Others improved with conservative treatment. The personal protective equipment of caregivers was not standard; despite the fact that only seven of the caregivers got COVID-19 during this period. However, it was not certain whether they got the infection from the hospital or outside. Fortunately, all of them improved without complications. The evidence available to date does not support a “child to adult” transmission of SARS Co V2.[17-18]

The mean duration of hospital stay was 5.53±1.58 days. No patient had hypoxemia or organ failure. Therefore, intensive care support was not required. This is due to the milder course of COVID-19 even with co-existing surgical emergencies. This finding supports the most published series of COVID-19 in children though serious infection and even death have also been reported in children.[3-6] This might be due to variable viral strains in this region or dissimilar immune responses to it.

Rescheduling of the non-emergent cases has already caused an enormous burden on the health care system. It is still not clear how long this critical period will go on. To reduce the sufferings of children with surgical needs, a gradual restart of scheduled surgeries is necessary considering the low transmissibility of COVID-19 in children.

## Conclusions

COVID-19 has made the world standstill. It is taking a toll on civilization. Fortunately, it is sparing the children so far but the impact on children’s surgery is already huge. Our study has shown the milder course of the disease even with a co-existing surgical emergency in children. The transmission of the disease from children to health care workers was also low. This might encourage the pediatric surgeons around the world for a gradual restart.

## Data Availability

All data are included in the manuscript and supplemental data. more specific inquiries will be entertained by the authors.

## Acknowledgement

In this desperate hour of our history, the heartiest gratitude goes to all those health workers, who are continually fighting risking their lives and saving thousands of lives while at peaked vulnerability.

## Contributors

**Dr. Md Samiul Hasan:** Conception, study design, data collection, data analysis, manuscript writing, and submission.

**Dr. Md Ayub Ali:** Obtaining ethical permission, Data collection, and manuscript revision.

**Dr. Umama Huq:** Data collection and manuscript editing.

## Funding statement

Authors received no fund for this article.

## Competing interest

No financial or nonfinancial benefits have been received or will be received from any party related directly or indirectly to the subject of this article.

## Ethical approval

Taken from Dhaka Shishu (Children) Hospital ethical committee.

## Abbreviations

COVID 19: coronavirus disease 19
SPSS: Statistical Package for the Social Science
IHPS: Infantile hypertrophic pyloric stenosis
ARM: Anorectal malformation
SARS CoV 2: Severe acute respiratory syndrome coronavirus 2
RT-PCR: Reverse transcription polymerase chain reaction
GI: Gastrointestinal

## Notes

### Competing Interest Statement

The authors have declared no competing interest.

### Funding Statement

The authors recieved no funding for the study.

### Author Declarations

Ethical permission was taken from hospital ethical review board. Approval ID: No. Admin/1050/2020/DSH

